# PRINCIPLE trial demonstrates scope for in-pandemic improvement in primary care antibiotic stewardship

**DOI:** 10.1101/2021.02.02.21250902

**Authors:** Simon de Lusignan, Mark Joy, Julian Sherlock, Manasa Tripathy, Oliver van Hecke, Oghenekome Gbinigie, John Williams, Christopher Butler, FD Richard Hobbs

## Abstract

**Background:** The Platform Randomised trial of INterventions against COVID-19 In older peoPLE (PRINCIPLE) trial has provided in-pandemic evidence of what does not work in the early primary care management of coronavirus-2019 disease (COVID-19). PRINCIPLE’s first finding was that azithromycin and doxycycline were not effective.

**Aim:** To explore the extent to which azithromycin and doxycycline were being used in-pandemic, and the scope for trial findings impacting on practice.

**Design and Setting:** We compared crude rates of prescribing and respiratory tract infections (RTI) in 2020, the pandemic year, with 2019, using the Oxford-Royal College of General Practitioners (RCGP) Research and Surveillance Centre (RSC).

**Methods:** We used a negative binomial model including age-band, gender, socioeconomic status, and NHS region to compare azithromycin and doxycycline lower respiratory tract infections (LRTI), upper respiratory tract infections (URTI), and influenza-like-illness (ILI) in 2020 with 2019; reporting incident rate ratios (IRR) between years and 95% confidence intervals (95%CI).

**Results:** Azithromycin prescriptions increased 7% in 2020 compared to 2019, whereas doxycycline decreased by 7%. Concurrently, LRTI and URTI incidence fell by over half (58.3% and 54.4% respectively) while ILI rose slightly (6.4%). The overall percentage of RTI prescribed azithromycin rose by 42.1% between 2019 and 2020, doxycycline increased by 33%.

Our adjusted IRR showed azithromycin prescribing was 22% higher in 2020 (IRR=1.22, 95%CI:1.19-1.26, p<0.0001), for every unit rise in confirmed COVID there was an associated 3% rise in prescription (IRR=1.026, 95%CI 1.024-1.0285, p<0.0001); whereas these measures were static for doxycycline.

**Conclusion:** PRINCIPLE trial flags scope for improvement in antimicrobial stewardship.

## Introduction

The Platform Randomised trial of INterventions against COVID-19 In older peoPLE (PRINCIPLE)^1^ has reported that two antibiotics, azithromycin and doxycycline, commonly prescribed to people with possible coronavirus 2019 infections (COVID-19) in primary care shows no meaningful benefit in terms of patient reported recovery outcomes.^2^ Azithromycin was included in PRINCIPLE between 23^rd^ May through to 30^th^ November 2020; doxycycline between 24^th^ July and 14^th^ December 2020.

However, there is no published evidence to know whether these antibiotics are being prescribed differently in primary care during the pandemic and therefore the extent to which these findings have potential to impact or change practice. Whilst National Institute for Health and Care Excellence (NICE) states: *As COVID-19 pneumonia is caused by a virus, antibiotics are ineffective*;^3^ it is entirely understandable given the disparities in who test positive^4^ and the excess mortality associated with COVID-19^5^ that general practitioners might have a lower threshold for prescribing antibiotics. NICE suggest that where antibiotics might be used doxycycline is the first choice.

We describe whether there was any change in the prescribing of these antibiotics in primary care during 2020, compared with the pre-pandemic year, taking into account any reduction in the incidence of consultations for respiratory tract infections (RTI) associated with the lockdowns, advice about where and when to seek help (e.g. NHS 111), and other non-pharmaceutical interventions (NPIs).^6^ We also report if azithromycin and doxycycline were more likely to be prescribed to people with a COVID-19 diagnosis.

## Method

### Data source and population

We used data from the Oxford-Royal College of General Practitioners (RCGP) Research and Surveillance Centre (RSC) sentinel network database. The RSC is one of Europe’s oldest sentinel systems and recruited to be nationally representative, with a history since 1967 of publishing a weekly report of monitored conditions – principally influenza-like-illness (ILI) and other respiratory infections and diseases.^7^ The RSC is actively involved, working closely with Public Health England (PHE) to deliver sentinel surveillance including of COVID-19.^8^

We carried out this analysis using sample of 397 general practices that met data quality standards in RSC with a total registered list size of 4,453,626. This subset was used because their antibiotic drug codes were carefully curated and met quality standards, and the patient group included also had near complete socioeconomic status recording (in most samples around 2.5% are missing).

General practices in the RSC are expected to code as a problem, in their computerised medical record, the diagnosis that the patient presents with. This is particularly so for our monitored conditions and key data required for determining vaccine effectiveness. We feedback to practices about their data quality via a dashboard, and do not include in our published reports those practices with monitored events where data quality level is below threshold.^9^

### Reporting crude differences

We described the monthly rate of prescribing of azithromycin and doxycycline comparing 2020 with 2019 (1/1/2020-31/12/2020 with 1/1/2019-31/12/2019). We next reported the monthly incidence of RTIs, again comparing 2020 with 2019. We only included coded cases which were labelled as an incident event or which were at least two weeks after any earlier recording. We compared lower respiratory infections (LRTI), upper respiratory infections (URTI), and influenza-like-illness (ILI) between the two years. We then summarised the rates of antibiotic use and of RTIs, providing 95% confidence intervals (95%CI) to discern differences in rates.

### Modelling the difference

We created negative binomial models; the first pair to report if there is any difference between the use of azithromycin and doxycycline in 2020, compared with 2019. We adjusted for age-band, gender, socioeconomic status using the index of multiple deprivation (derived from post code), NHS region (though we combined midlands and east regions to provide a more balanced distribution), and the incidence of LRTI, URTI and ILI. Concerning age, the population included in PRINCIPLE was people over 50 years old with one or more comorbidities and population age 65 years or older. We therefore included a 65 years and older age-band in our analysis as most readily compared with PRINCIPLE, but decided to include antibiotic use across all age-bands. We report an incident rate ratio (IRR) and 95% CI, and significance. We ran the same model using 2020 data to explore whether COVID-19 cases were more likely prescribed azithromycin or doxycycline.

### Sensitivity analysis

We conducted a sensitivity analysis to explore whether the RSC practices had higher rates of prescription of antibiotics that the rest of England. We did this because the PRINCIPLE trial has been widely promoted to RSC practices and this may have encouraged increased prescribing. We therefore compared the pattern of prescribing in the RSC using national data from OpenPrescribing.^10^ We also compared the lowest prescribing rates in 2020, and average monthly rates of prescribing to see if there were any difference in prescribing rates. As OpenPrescribing does not publish a denominator we use the list size of all English practices published by NHS Digital as denominator.^11^ Finally, we reported from OpenPrescribing the mean difference in prescribing of these antibiotics between 2020 and 2019, comparing the first 10 months of the year, as there were only data to October 2020 available.

### Ethical considerations

This investigation used pseudonymised data held by the Oxford-RCGP RSC sentinel network. The investigation is classified by the Health Research Authority Decision tool (http://www.hra-decisiontools.org.uk/research/) as not being considered research, and not requiring formal research ethics approval. This investigation was approved by the Oxford-RCGP RSC Joint Research and Surveillance Centre Committee.

## Results

### Crude rates of azithromycin and doxycycline prescribing

Overall, the number of azithromycin prescriptions increased by 6.98% between 2019 and 2020, while that of doxycycline fell by 7.02% (Supplementary file, Table S4.1). However, the pattern of prescriptions of both antibiotics through 2020 were very different from 2019, the latter followed a more typical pattern.

In January and February 2020 prescriptions of azithromycin and doxycycline were similar to those in 2019. However, in March 2020 both antibiotics were prescribed much more. This was coincident with the first wave of COVID-19. From then on, the use of these antibiotics in 2020 diverged in their use. Azithromycin was prescribed in 2020 at or above the level prescribed in 2019, the converse was true for doxycycline (Figure 1, tables of rates S1.1 and S1.2 in Supplementary file).

**Figure 1:**
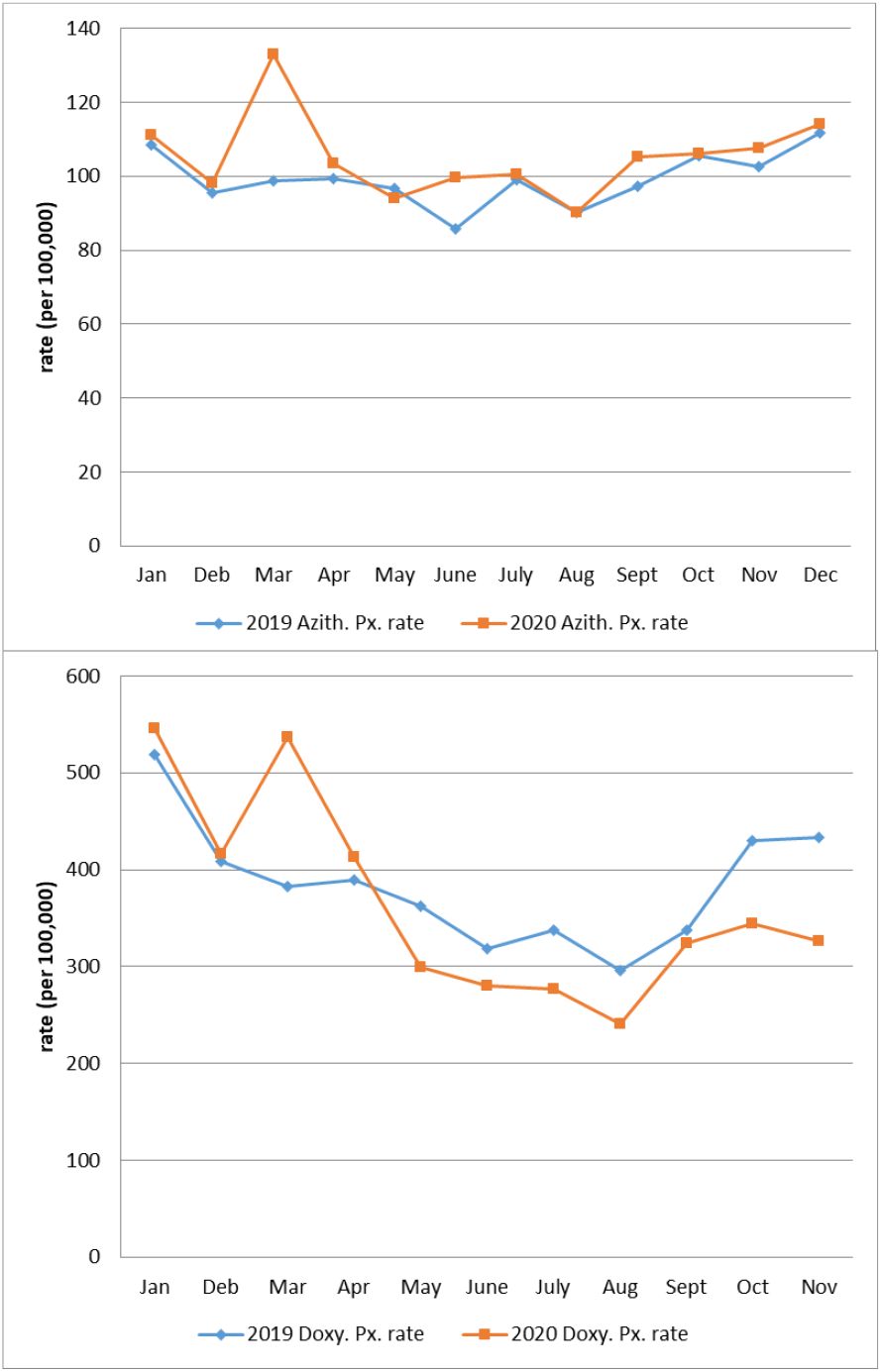
Prescribing of azithromycin (top) and doxycycline (bottom) by month within the RSC. 2020 prescription of both antibiotics (red line) was very similar in January and February to 2019 rates (blue line), then in March there was a peak of prescribing in 2020 coincident with the first wave of the COVID-19 pandemic. Thereafter azithromycin was prescribed in 2020 at or above the level in 2019, whereas doxycycline was prescribed less.

The monthly incidence of patient consultations with a RTI, diagnosed by GPs and recorded in their computerised medical records, was over 50% lower in 2020 than in 2019. For LRTI and URTI the incidence was lower every month. For example, the year 2019 shows a typical pattern for LRTI and URTI; the incidence is lowest at the end of the school summer holidays, peaks at the end of the year, and then falls slowly back to its low point at the end of August (Figure 1). In contrast, the pattern of ILI is different each year. Its peak generally follows the peak in influenza A. For example, 2019 was a typical year with a peak incidence around or just after the year end. In comparison, 2020 showed changes in ILI incidence largely reflecting the waves of the COVID-19 pandemic with peaks in March, April coinciding with the first wave, and a second rise as schools go back, with this modestly increased level continuing to year end (Figure 2, Supplementary file Table S2.3).

**Figure 2:**
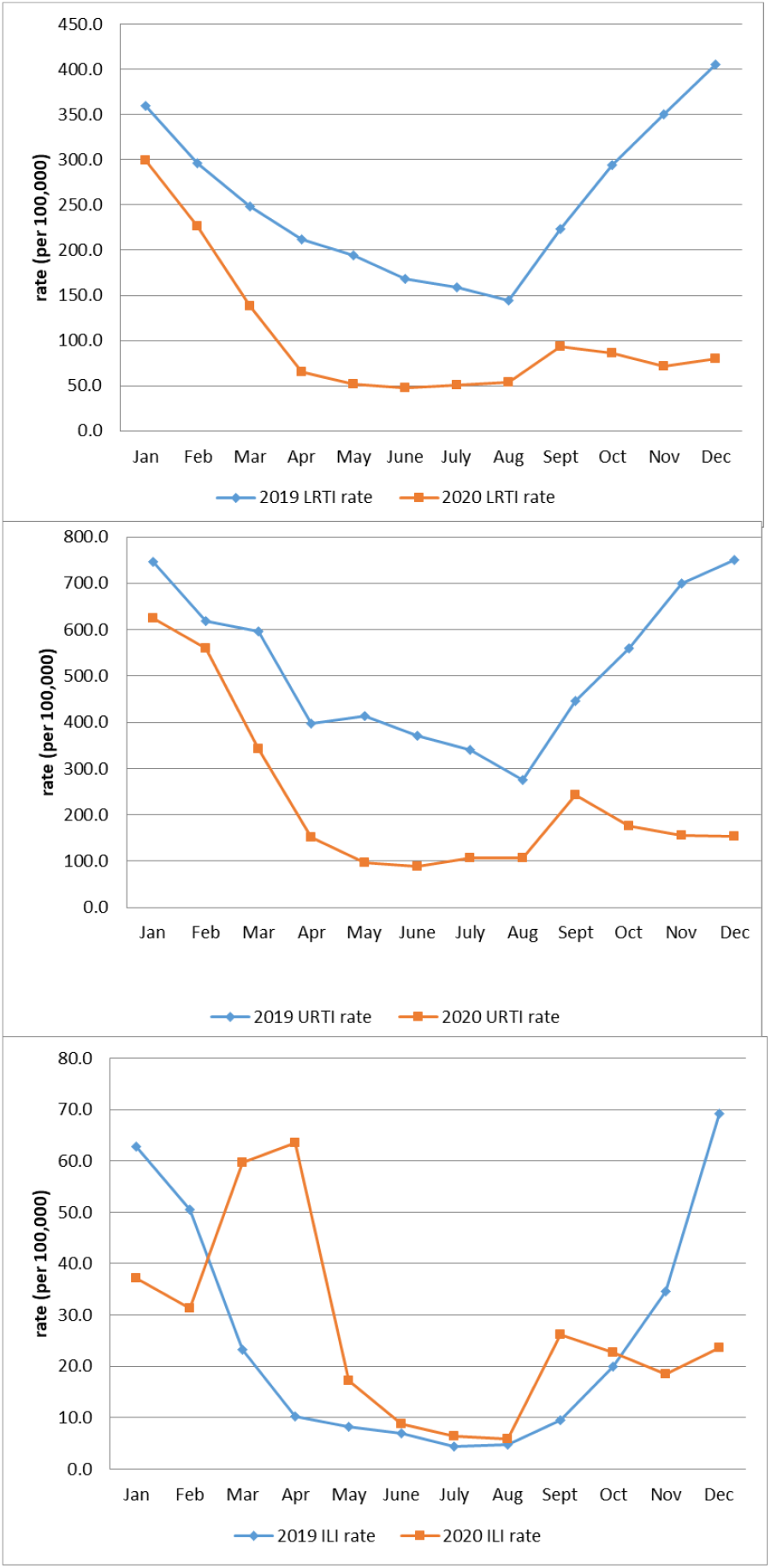
Comparison of monthly incidence of consultations for LRTI (top), URTI (middle) and ILI (bottom) comparing 2020 (red) with 2019 (blue) in the RSC dataset. There was a lower incidence of LRTI and URI in 2020 compared with 2019, with a small peak when schools returned in September 2020. ILI peaked with the first wave of the COVID-19 pandemic also with the return to school.

Unsurprisingly, with social distancing measures and lockdown, there was a lower rate of LRTI (−58.3%) and URTI (−54.4%) consultations across all age bands. ILI consultation rates overall were slightly increased (6.4%) between the years. However, there was a fall in ILI in males under 16 years old and a rise in presentation of females age 16 to 64 years in 2020 compared with 2019 (Table 1).

**Table 1:** Comparison of rates of prescription of doxycycline and azithromycin in 2020 with 2019. In people 65 years old and older there was a decrease in doxycycline use but an increase in azithromycin prescription. LRTI and URTI incidence fell across all age bands and both genders. ILI was much more similar between years.

|  |  |  | 2019 |  | 2020 |  |
| --- | --- | --- | --- | --- | --- | --- |
|  |  | Age | Female | Male | Female | Male |
| Antibiotic Rates<br>(per 100,000) | Doxy. | under 16 | 21.74 (20.4,23.2)* | 17.75 (16.5,19.0) | 19.12 (17.8,20.5) | 15.96 (14.8,17.2) |
|  |  | 16-64 | 385.28 (382.1,388.5) | 231.99 (229.6,234.4) | 380.15 (377.8,383.3) | 222.57 (220.2,224.9) |
|  |  | 65+ | 1136.15 (1126.4,1145.9) | 1038.78 (1028.6,1048.9) | 968.28 (959.3,977.3) | 913.57 (904.1,923.1) |
|  | Azith. | under 16 | 56.89 (54.6,59.2) | 67.50 (65.1,69.9) | 53.29 (51.1,55.5) | 75.91 (73.4,78.5) |
|  |  | 16-64 | 70.02 (68.7,71.4) | 39.39 (38.4,40.4) | 70.69 (69.4,72.1) | 40.77 (39.8,41.8) |
|  |  | 65+ | 305.17 (300.1,310.3) | 288.43 (283.1,293.9) | 339.13 (333.8,344.5) | 307.59 (302.1,313.2) |
| Resp. Disease<br>Rates (per<br>100,000) | LRTI | under 16 | 229.25 (224.67,233.82) | 292.18 (287.1,297.2) | 68.43 (65.9,70.9) | 90.9 (88.1,93.8) |
|  |  | 16-64 | 191.30 (189.1, 193.54) | 126.84 (125.0,128.6) | 81.45 (79.9,82.9) | 53.2 (52.0,54.4) |
|  |  | 65+ | 609.27 (602.05,616.5) | 568.32 (560.8,575.8) | 267.9 (263.2,272.8) | 268.0 (262.9,273.2) |
|  | URTI | under 16 | 1320.33 (1309.5,1331.3) | 1349.86 (1339.1,1360.7) | 485.15 (455.0,467.7) | 493.1 (486.6,499.7) |
|  |  | 16-64 | 485.65 (482.1,489.2) | 229.3 (226.9,231.8) | 265.73 (254.9,260.1) | 117.38 (115.7,119.1) |
|  |  | 65+ | 285.17 (280.3,290.1) | 208.6 (204.1,213.2) | 148.01 (144.5,151.6) | 104.61 (101.4,107.9) |
|  | ILI | under 16 | 19.30 (17.1,19.7) | 20.78 (19.5,22.2) | 16.38 (15.2,17.7) | 16.23 (15.1,17.5) |
|  |  | 16-64 | 32.57 (30.8,32.6) | 22.15 (21.4,22.9) | 37.68 (36.7,38.7) | 21.86 (21.1,22.6) |
|  |  | 65+ | 25.40 (28.2,31.7) | 22.18 (20.7,23.7) | 29.41 (27.9,31.0) | 24.38 (22.9,25.9) |

In 2020, the use of azithromycin increased in both genders in the 65 years and over age-band, and in males under 16 years old, compared with 2019, the overall increase was 6.98%. Not-withstanding the observed fall in RTI rates, there has been a concomitant rise of 0.21% (95%CI: 0.16-0.26), p<0.0001) in the proportion of respiratory infections where azithromycin was prescribed, a 42.1% rise on the previous year.

The converse occurred with doxycycline with a reduction in the rate of prescribing in the 65 years and older age-band. Overall, there was a decline in prescribing of 7.02% between 2019 and 2020. However, in relation to the fall in acute respiratory infections in 2020, there was nearly a 4% rise (3.93%, 95%CI:[3.73-4.14], *p*<0.0001) of doxycycline prescribing, a 33.1% rise on the previous year.

After having adjusted for age, gender, socioeconomic status, NHS region and RTIs (LRTI, URTI and ILI), the negative binomial model indicated that the frequency of azithromycin prescriptions (for any reason) in the RSC registered population is 22% higher in 2020 compared to 2019 (IRR=1.22, 95%CI 1.19-1.26, p<0.0001, Table 2).

**Table 2:**
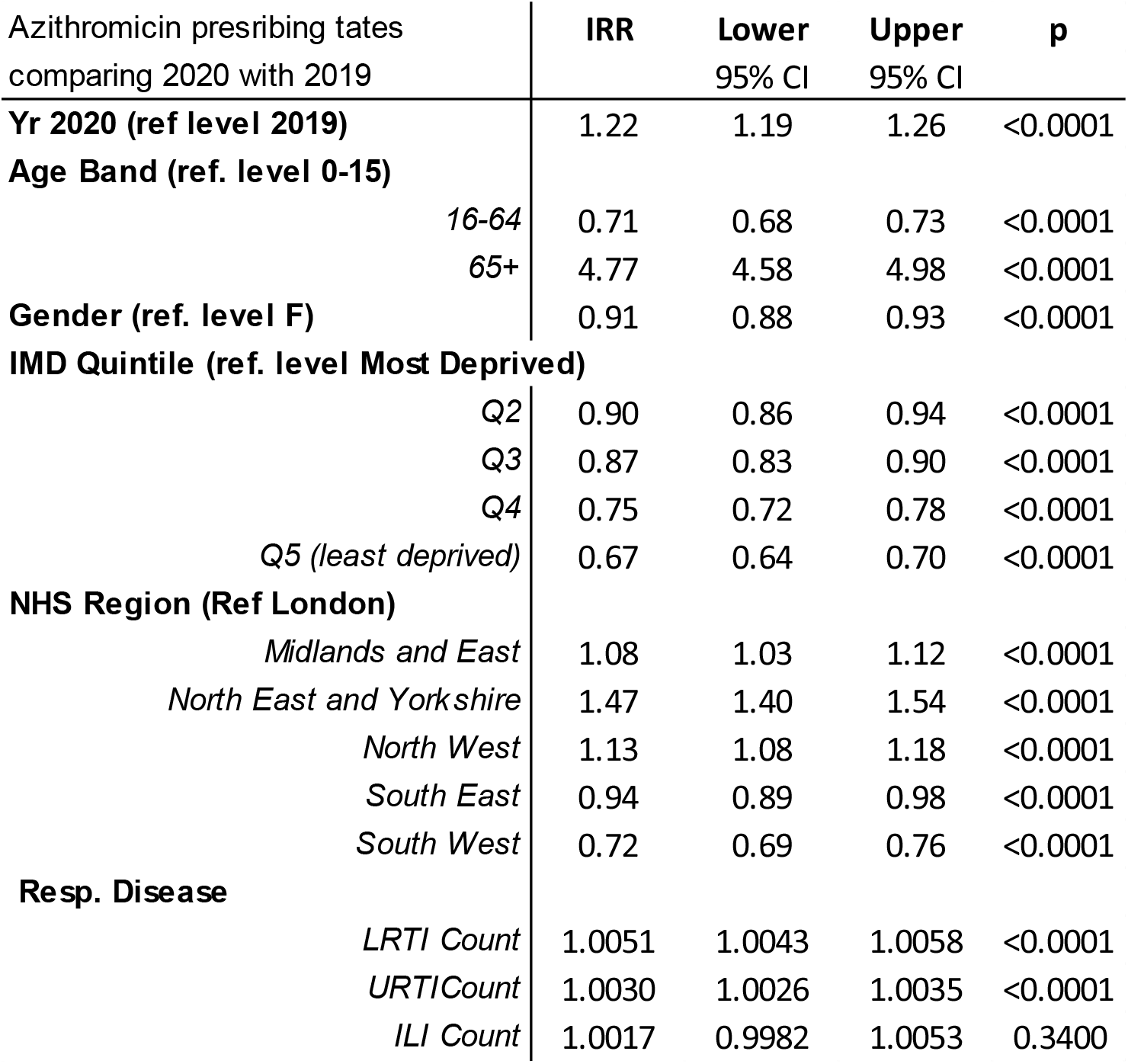
Model reporting the incident rate ratio (IRR) comparing prescribing of azithromycin in 2020 with 2019. Taking the variables in the model into account there was a 22% increase, with people 65 years and older, female gender, the most deprived, northern regions and people with LRTI and URTI.

| Azithromycin prescribing rates<br>comparing 2020 with 2019 |  | IRR | Lower<br>95% CI | Upper<br>95% CI | p |
| --- | --- | --- | --- | --- | --- |
| **Yr 2020 (ref level 2019)** |  | 1.22 | 1.19 | 1.26 | <0.0001 |
| **Age Band (ref. level 0-15)** |  |  |  |  |  |
|  | 16-64 | 0.71 | 0.68 | 0.73 | <0.0001 |
|  | 65+ | 4.77 | 4.58 | 4.98 | <0.0001 |
| **Gender (ref. level F)** |  | 0.91 | 0.88 | 0.93 | <0.0001 |
| **IMD Quintile (ref. level Most Deprived)** |  |  |  |  |  |
|  | Q2 | 0.90 | 0.86 | 0.94 | <0.0001 |
|  | Q3 | 0.87 | 0.83 | 0.90 | <0.0001 |
|  | Q4 | 0.75 | 0.72 | 0.78 | <0.0001 |
|  | Q5 (least deprived) | 0.67 | 0.64 | 0.70 | <0.0001 |
| **NHS Region (Ref London)** |  |  |  |  |  |
|  | Midlands and East | 1.08 | 1.03 | 1.12 | <0.0001 |
|  | North East and Yorkshire | 1.47 | 1.40 | 1.54 | <0.0001 |
|  | North West | 1.13 | 1.08 | 1.18 | <0.0001 |
|  | South East | 0.94 | 0.89 | 0.98 | <0.0001 |
|  | South West | 0.72 | 0.69 | 0.76 | <0.0001 |
| **Resp. Disease** |  |  |  |  |  |
|  | LRTI Count | 1.0051 | 1.0043 | 1.0058 | <0.0001 |
|  | URTI Count | 1.0030 | 1.0026 | 1.0035 | <0.0001 |
|  | ILI Count | 1.0017 | 0.9982 | 1.0053 | 0.3400 |

In addition, for every unit rise in COVID confirmed status there was an associated 3% rise in azithromycin prescription (IRR=1.026, 95%CI 1.024-1.0285, p<0.0001, Table 3). With azithromycin, there was a much higher rate of prescribing to those age 65 years and over, and less to those age 16 to 64 years. There was less azithromycin prescribing for males compared with females, higher rates of prescribing to the most deprived regions and in the north compared to the south. Comparing 2020 with 2019 overall there was more azithromycin prescribing for people with LRTI and URTI. During 2020 there was a higher rate of prescribing to people with LRTI and ILI and reduced prescribing to people with URTI.

**Table 3:** Azithromycin prescribing in cases of COVID-19, for each unit rise in COVID-19 cases there has been a 3% rise in azithromycin prescriptions. Age 65 years and older, female gender, being more deprived, northern regions LRTI or ILI infections are all associated with a higher rate of prescribing

| Azithromycin prescribing rate | IRR | Lower<br>95% CI | Upper<br>95% CI | p |
| --- | --- | --- | --- | --- |
| **Covid19 Confirmed Count** | 1.03 | 1.02 | 1.03 | <0.0001 |
| **Age Band (ref. level 0-15)** |  |  |  |  |
| 16-64 | 0.25 | 0.20 | 0.31 | <0.0001 |
| 65+ | 10.95 | 8.67 | 13.83 | <0.0001 |
| **Gender (ref. level F)** | 0.54 | 0.45 | 0.65 | <0.0001 |
| **IMD Quintile (ref. level Most Deprived)** |  |  |  |  |
| Q2 | 0.54 | 0.41 | 0.72 | <0.0001 |
| Q3 | 0.41 | 0.31 | 0.55 | <0.0001 |
| Q4 | 0.54 | 0.41 | 0.72 | <0.0001 |
| Q5 (least deprived) | 0.66 | 0.50 | 0.88 | 0.0048 |
| **NHS Region (Ref London)** |  |  |  |  |
| Midlands and East | 5.73 | 4.28 | 7.69 | <0.0001 |
| North East and Yorkshire | 12.88 | 9.18 | 18.07 | <0.0001 |
| North West | 10.31 | 7.34 | 14.49 | <0.0001 |
| South East | 2.82 | 2.01 | 3.96 | <0.0001 |
| South West | 1.41 | 1.01 | 1.98 | 0.0453 |
| **Resp. Disease** |  |  |  |  |
| LRTI Count | 1.94 | 1.92 | 1.97 | <0.0001 |
| URTI Count | 0.89 | 0.88 | 0.90 | <0.0001 |
| ILI Count | 1.60 | 1.54 | 1.68 | <0.0001 |

For doxycycline, there was no change in the rate of prescribing comparing 2020 with 2019 (IRR=1.012, 95%CI 0.994-1;030, p=0.199). Female gender, the most deprived quintile, midlands and southwest region LRTI and ILI were associated with higher rates of prescription in 2020 compared with 2019 (see supplementary material table S3.1).

Adjusting for age, gender, socioeconomic status, region and RTI, there was a very small rise of 0.3% in the rate of prescribing in doxycycline (IRR=1.0003, 95%CI 1.0002-1;0005, p<0.0001). Female gender, the most deprived quintile, midlands and southwest region LRTI and ILI were associated with higher rates of prescription of doxycycline (see supplementary material table S3.2).

Our sensitivity analysis showed a very similar pattern of month-by-month azithromycin and doxycycline prescribing as OpenPrescribing (Figure 3), a national database. We used the NHS Digital registered patient population denominator (N=60,570,367); compared with the RSC sample denominator (N=4,453,626). We compared the rates of the lowest monthly overall prescription count in 2020. We found a prescribing rate (per registered patient) of 0.248% for OpenPrescribing and 0.225% for the RSC for doxycycline; and 0.099% and 0.075% for azithromycin. We also compared the year-on-year change. In OpenPrescribing azithromycin prescribing increased by 7.25% and doxycycline reduced by 2.31%, between 2019 and 2010, compared with 6.98% and =7.02% in the RSC for azithromycin and doxycycline respectively (Supplementary file Table S4.1).

**Figure 3:**
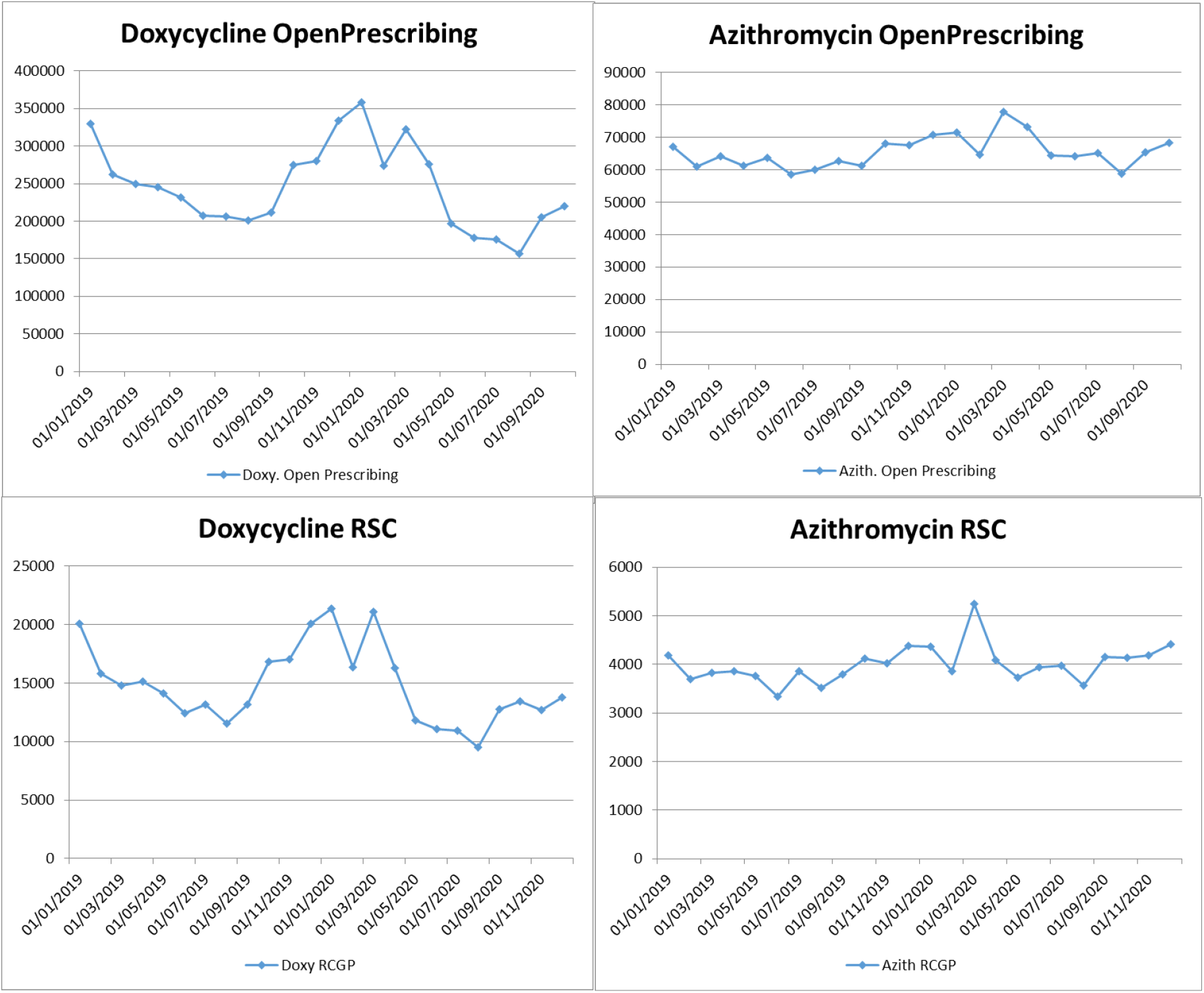
Monthly pattern of doxycycline and azithromycin, prescription counts, for 2019 and 2020 (OpenPrescribing data are only available up to October 2020).

## Discussion

### Main findings

Azithromycin prescribing, for all indications, increased by 7% in 2020 compared to 2019; doxycycline prescribing reduced by the same amount (7%). The RSC mirrors the pattern and rate of azithromycin prescribing reported by OpenPrescribing, whilst there was a similar pattern of doxycycline prescription OpenPrescribing reports a smaller year-on-year decline.

Prescribing of both antibiotics peaked in the first wave of COVID-19, in March 2020, with the use of azithromycin prescribing overall staying like last year, while there is a lower rate of doxycycline prescribing overall. There was no equivalent peak of prescribing these antibiotics in the second wave of COVID-19 in the autumn.

Through 2020 there has been a much lower incidence of consultations coded with a diagnosis of upper and lower RTIs, though ILI incidence increased has been more complex. ILI incidence has mirrored COVID-19 infection.

The rate of azithromycin prescribing, taking into account the other variables in our model, has increased by 22% in 2020 compared to 2019. Further, as the number of COVID-19 cases increased, so the prescribing of azithromycin increased. Doxycycline, shows a different pattern. Its rate of prescription has not changed between 2019 and 2020, although there has been an increase in prescriptions of around 4% for RTIs and its use marginally changes with increasing COVID-19 cases.

These observations are unsurprising given an understandable imperative in clinical practice to trial interventions that might improve COVID-19 and are in the public eye. Furthermore, more remote consultations have increased substantially,^12^ and may make it harder for clinicians to accurately diagnose pneumonia that has complicated COVID-19, leading to a reduced threshold to prescribe antibiotics. However, observational studies suggest that even amongst patients hospitalised with COVID-19, the prevalence of bacterial super-infection is low with estimates of around 3.5%.^13,14^ Routine antimicrobial prescription for COVID-19 patients in the community should now change in light of the new evidence.

### Implications of the findings

The PRINCIPLE trial has demonstrated that there is no benefit from either antibiotic in early treatment of COVID-19 in the absence of bacterial pneumonia. There is therefore considerable scope to reduce the prescribing of these antibiotics in primary care.

Additionally, it appears that awareness of, or involvement in the PRINCIPLE trial by RSC practices does not seem to be associated with higher rates of prescribing than those seen in national data reported by OpenPrescribing.

### Strengths and limitations

The strength of this analysis is that the RSC has good data quality and is able to capture routine data about RTIs and their incidence. The network encourages our key monitored conditions to be coded as problems with an episode type, to distinguish new from follow-up consultations. Where episode type is not recorded, we use a time interval to distinguish incidence from follow-up cases.^15^ RTI data are extracted from practices either daily or twice weekly, with data fed back to practices via a dashboard to try to raise awareness of data quality.^16^

Comparing the use between years of azithromycin and doxycycline is complex. Firstly, azithromycin prescribing has risen, but the same proportion by which doxycycline has fallen (7%) between 2019 and 2020. However, the incidence in the respiratory conditions they treat has fallen by a greater proportion. Both antibiotics have had a significantly increased use in respiratory infections, with the NICE guidance, which suggests using doxycycline first line, not seeming to be associated with an overall increase in use.^3^

A larger proportion of people who consult with a respiratory infection were prescribed these antibiotics in 2020, compared with 2019. However, the situation is further complicated by the first two waves of the COVID-19 pandemic affecting England during 2020, with considerable uncertainty as to what medications may be of benefit. Finally, only people aged 50years and above were eligible for the PRINCIPLE trial, these data are from across all ages.

## Conclusions

An in-pandemic trial has studied the value of prescribing two antibiotics for people with COVID-19. Given the lack of efficacy of azithromycin and doxycycline demonstrated in the PRINCIPLE trial we should apply good antibiotic stewardship and use them less. The national sentinel network, the Oxford-RCGP RSC has demonstrated that these antibiotics are being extensively used and that they are being prescribed to a higher proportion of people with respiratory infections then in the pre-pandemic year. There is scope, in-pandemic, to reduce the use of doxycycline and azithromycin in primary care.

## Data Availability

The RCGP RSC dataset can be accessed by researchers, approval is on a project-by-project basis (www.rcgp.org.uk/rsc). Ethical approval by an NHS Research Ethics Committee is needed before any data release/other appropriate approval. Researchers wishing to directly analyse patient-level pseudonymised data will be required to complete information governance training and work on the data from the secure servers at the University of Oxford. Patient-level data cannot be taken out of the secure network.

## Acknowledgements

Patients and practices in the Oxford-RCGP RSC who allow data sharing. UKRI for its support of the PRINCIPLE trial. EMIS, TPP, In-Practice Systems and Wellbeing software for their collaboration with pseudonymised data extraction. RCGP and NIHR CRN for their support in encouraging practice involvement with the Oxford-RCGP RSC and the PRINCIPLE trial. FDRH acknowledges part support from the NIHR School for Primary Care Research (SPCR), the NIHR Applied Research Collaboration (ARC) Oxford, and the NIHR Oxford BRC. Public Health England (PHE) for their support with participant testing of COVID-19.

## Declaration of interest

CB and FDRH are co-Principal and SdeL an investigator of the PRINCIPLE trial. SdeL is Director of the Oxford-RCGP RSC.

## Notes

### Competing Interest Statement

Christopher Butler (CB) and FD Richard Hobbs (FDRH) are co-Principal and Simon de Lusignan (SdeL) an investigator of the PRINCIPLE trial. SdeL is Director of the Oxford-RCGP RSC.

### Clinical Trial

ISRCTN86534580

### Clinical Protocols

https://publichealth.jmir.org/2020/3/e19773/

### Funding Statement

PRINCIPLE is funded by UK Research and Innovation and the Department of Health and Social Care through the National Institute for Health Research. FD Richard Hobbs acknowledges part support from the NIHR School for Primary Care Research (SPCR), the NIHR Applied Research Collaboration (ARC) Oxford, and the NIHR Oxford BRC.

## References

1 ISRCTN Registry. PRINCIPLE: A trial evaluating treatments for suspected COVID-19 in people aged 50 years and above with pre-existing conditions and those aged 65 years and above. ISRCTN86534580 https://doi.org/10.1186/ISRCTN86534580

2 Butler CC. et al., University of Oxford (2021) Azithromycin and doxycycline are not generally effective against COVID-19 in patients treated at home, shows PRINCIPLE trial [Press Release]. 25 January. Available at: https://www.principletrial.org/news/azithromycin-and-doxycycline-are-not-generally-effective-treatments-for-covid-19-shows-principle-trial (Accessed: 01 February 2021)

3 National Institute for Health and Care Excellence (NICE). COVID-19 rapid guideline: managing suspected or confirmed pneumonia in adults in the community. NICE guideline [NG165] Published: 03 April 2020 Last updated: 23 April 2020. URL: https://www.nice.org.uk/guidance/ng165/chapter/4-Managing-suspected-or-confirmed-pneumonia#antibiotic-treatment

4 de Lusignan S, Dorward J, Correa A, Jones N, Akinyemi O, Amirthalingam G, Andrews N, Byford R, Dabrera G, Elliot A, Ellis J, Ferreira F, Lopez Bernal J, Okusi C, Ramsay M, Sherlock J, Smith G, Williams J, Howsam G, Zambon M, Joy M, Hobbs FDR. Risk factors for SARS-CoV-2 among patients in the Oxford Royal College of General Practitioners Research and Surveillance Centre primary care network: a cross-sectional study. Lancet Infect Dis. 2020 Sep;20(9):1034–1042. doi: 10.1016/S1473-3099(20)30371-6.

5 Joy M, Hobbs FR, Bernal JL, Sherlock J, Amirthalingam G, McGagh D, Akinyemi O, Byford R, Dabrera G, Dorward J, Ellis J, Ferreira F, Jones N, Oke J, Okusi C, Nicholson BD, Ramsay M, Sheppard JP, Sinnathamby M, Zambon M, Howsam G, Williams J, de Lusignan S. Excess mortality in the first COVID pandemic peak: cross-sectional analyses of the impact of age, sex, ethnicity, household size, and long-term conditions in people of known SARS-CoV-2 status in England. Br J Gen Pract. 2020 Nov 26;70(701):e890–e898. doi: 10.3399/bjgp20X713393.

6 Flaxman S, Mishra S, Gandy A, Unwin HJT, Mellan TA, Coupland H, Whittaker C, Zhu H, Berah T, Eaton JW, Monod M; Imperial College COVID-19 Response Team, Ghani AC, Donnelly CA, Riley S, Vollmer MAC, Ferguson NM, Okell LC, Bhatt S. Estimating the effects of non-pharmaceutical interventions on COVID-19 in Europe. Nature. 2020 Aug;584(7820):257–261. doi: 10.1038/s41586-020-2405-7.

7 de Lusignan S, Correa A, Smith GE, Yonova I, Pebody R, Ferreira F, Elliot AJ, Fleming D. RCGP Research and Surveillance Centre: 50 years’ surveillance of influenza, infections, and respiratory conditions. Br J Gen Pract. 2017 Oct;67(663):440–441. doi: 10.3399/bjgp17X692645.

8 de Lusignan S, Lopez Bernal J, Byford R, Amirthalingam G, Ferreira F, Akinyemi O, Andrews N, Campbell H, Dabrera G, Deeks A, Elliot A, Krajenbrink E, Liyanage H, Mcgagh D, Okusi C, Parimalanathan V, Ramsay M, Smith G, Tripathy M, Williams J, Victor W, Zambon M, Howsam G, Nicholson B, Tzortziou Brown V, Butler C, Joy M, Hobbs R. Seasonal influenza surveillance and vaccine effectiveness at a time of co-circulating COVID-19: Oxford-Royal College of General Practitioners (RCGP) Research and Surveillance Centre (RSC) and Public Health England (PHE) protocol for winter 2020/21. JMIR Public Health and Surveillance. 08/12/2020:24341 (forthcoming/in press). doi: 10.2196/24341 URL: https://preprints.jmir.org/preprint/24341

9 Liyanage H, Akinyemi O, Pathirannahelage S, Joy M, de Lusignan S. Near Real Time Feedback of Seasonal Influenza Vaccination and Virological Sampling: Dashboard Utilisation in a Primary Care Sentinel Network. Stud Health Technol Inform. 2020 Jun 16;270:1339–1340. doi: 10.3233/SHTI200431.

10 Curtis HJ, Goldacre B. OpenPrescribing: normalised data and software tool to research trends in English NHS primary care prescribing 1998-2016. BMJ Open. 2018 Feb 23;8(2):e019921. doi: 10.1136/bmjopen-2017-019921.

11 NHS Digital. Patients Registered at a GP Practice December 2020. URL: https://digital.nhs.uk/data-and-information/publications/statistical/patients-registered-at-a-gp-practice/december-2020

12 Joy M, McGagh D, Jones N, Liyanage H, Sherlock J, Parimalanathan V, Akinyemi O, van Vlymen J, Howsam G, Marshall M, Hobbs FR, de Lusignan S. Reorganisation of primary care for older adults during COVID-19: a cross-sectional database study in the UK. Br J Gen Pract. 2020 Jul 30;70(697):e540–e547. doi: 10.3399/bjgp20X710933.

13 Vaughn, V.M., Gandhi, T., Petty, L.A., Patel, P.K., Prescott, H.C., Malani, A.N., Ratz, D., McLaughlin, E., Chopra, V. and Flanders, S.A., 2020. Empiric Antibacterial Therapy and Community-onset Bacterial Co-infection in Patients Hospitalized with COVID-19: A Multi-Hospital Cohort Study. Clinical infectious diseases: an official publication of the Infectious Diseases Society of America.

14 Nori, P., Cowman, K., Chen, V., Bartash, R., Szymczak, W., Madaline, T., Katiyar, C.P., Jain, R., Aldrich, M., Weston, G. and Gialanella, P., 2021. Bacterial and fungal coinfections in COVID-19 patients hospitalized during the New York City pandemic surge. Infection Control & Hospital Epidemiology, 42(1), pp.84–88.

15 Smith N, Livina V, Byford R, Ferreira F, Yonova I, de Lusignan S. Automated Differentiation of Incident and Prevalent Cases in Primary Care Computerised Medical Records (CMR). Stud Health Technol Inform. 2018;247:151–155.

16 Pathirannehelage S, Kumarapeli P, Byford R, Yonova I, Ferreira F, de Lusignan S. Uptake of a Dashboard Designed to Give Realtime Feedback to a Sentinel Network About Key Data Required for Influenza Vaccine Effectiveness Studies. Stud Health Technol Inform. 2018;247:161–165.

