## Supplementary File for "PRINCIPLE trial demonstrates scope for in-pandemic improvement in primary care antibiotic stewardship"

**Antibiotic prescribing:**

Tables S1.1 and S1.2 shows the monthly prescribing rate of azithromycin and doxycycline in 2019 and 2020. These data are used for Figure 1.

| Month | 2019 Azith. Px. rate | 2020 Azith. Px. rate |
|---|---|---|
| Jan | 108.4 | 111.1 |
| Deb | 95.5 | 98.0 |
| Mar | 98.7 | 133.0 |
| Apr | 99.2 | 103.5 |
| May | 96.8 | 94.2 |
| June | 85.9 | 99.7 |
| July | 99.0 | 100.6 |
| Aug | 90.3 | 90.1 |
| Sept | 97.1 | 105.4 |
| Oct | 105.5 | 106.0 |
| Nov | 102.5 | 107.6 |
| Dec | 111.7 | 114.1 |

**Table S1.1 Azithromycin prescription rate by month per 100,000 registered patients for 2019 and 2020**

| Month | 2019 Doxy. Px. rate | 2020 Doxy. Px. rate |
|---|---|---|
| Jan | 519.0 | 545.6 |
| Deb | 408.6 | 416.7 |
| Mar | 382.4 | 536.4 |
| Apr | 389.4 | 412.5 |
| May | 362.7 | 298.8 |
| June | 318.9 | 279.7 |
| July | 337.9 | 276.7 |
| Aug | 296.1 | 240.6 |
| Sept | 337.2 | 324.2 |
| Oct | 429.8 | 344.7 |
| Nov | 433.8 | 326.6 |
| Dec | 511.9 | 357.1 |

**Table S1.2 Doxycycline prescription rate by month per 100,000 registered patients for 2019 and 2020**

**Monthly rates of respiratory tract infections (RTI) comparing 2019 with 2020:**
These are the monthly rates of LRTI, URTI and ILI for 2019 and 2020, rates per 10,000 registered populations (Tables S2.1 to S2.3). These data were used to support Figure 2.

| Month | 2019 LRTI rate | 2020 LRTI rate |
|---|---|---|
| Jan | 359.4 | 299.4 |
| Feb | 296.1 | 226.7 |
| Mar | 248.4 | 137.9 |
| Apr | 211.7 | 65.1 |
| May | 194.4 | 51.5 |
| June | 167.8 | 48.0 |
| July | 159.2 | 50.3 |
| Aug | 143.9 | 53.8 |
| Sept | 223.1 | 93.7 |
| Oct | 294.4 | 86.3 |
| Nov | 350.2 | 71.7 |
| Dec | 405.0 | 80.3 |

**Table S2.1: LRTI rates, monthly incidence comparing 2019 and 2020, per 10,000 registered patients**

| Month | 2019 URTI rate | 2020 URTI rate |
|---|---|---|
| Jan | 747.2 | 624.4 |
| Feb | 618.5 | 560.3 |
| Mar | 595.7 | 341.7 |
| Apr | 397.4 | 151.3 |
| May | 413.9 | 97.6 |
| June | 369.9 | 88.9 |
| July | 340.6 | 106.6 |
| Aug | 275.5 | 107.8 |
| Sept | 445.0 | 243.0 |
| Oct | 559.9 | 176.2 |
| Nov | 700.0 | 156.5 |
| Dec | 751.0 | 154.0 |

**Table S2.2: URTI rates, monthly incidence comparing 2019 and 2020, per 10,000 registered patients**

| Month | 2019 ILI rate | 2020 ILI rate |
|---|---|---|
| Jan | 62.8 | 37.2 |
| Feb | 50.6 | 31.3 |
| Mar | 23.3 | 59.6 |
| Apr | 10.3 | 63.6 |
| May | 8.2 | 17.3 |
| June | 6.9 | 8.7 |
| July | 4.4 | 6.5 |
| Aug | 4.7 | 5.8 |
| Sept | 9.5 | 26.1 |
| Oct | 19.9 | 22.7 |
| Nov | 34.5 | 18.5 |
| Dec | 69.1 | 23.7 |

**Table S2.3 ILI rates, monthly incidence comparing 2019 and 2020, per 10,000 registered patients**

**Change in respiratory infections 2020 compared to 2019:**
We cross-tabulated the change in respiratory infections between 2020 and 2019, by RTI, showing the percentage change (Table S3.1). Overall RTI reduced over 50%, but there was a small rise in ILI.

| | Resp. Tract Infection Consultations in RSC | | % Change |
|---|---|---|---|
| | **2019** | **2020** | |
| **LRTI** | 118,940 | 49,631 | -58.3 |
| **URTI** | 241,997 | 110,230 | -54.4 |
| **ILI** | 11,846 | 12,607 | 6.4 |
| **Total** | **372,783** | **172,468** | **-53.7** |

**Table S3.1: Respiratory tract infections recorded in the RSC 2019 and 2020, and percentage change**

We next show (Tables S3.2 and S3.3) the change in prescribing of azithromycin and doxycycline comparing the RTI consultations where these antibiotics were prescribed with those where they were not.

| | All RTI Consultations in RSC | | % Change |
|---|---|---|---|
| | **2019** | **2020** | *p<0.0001* |
| RTI no azithromycin Px | 370885 | 171220 | -53.8 |
| RTI with azithromycin Px | 1898 | 1248 | -34.2 |
| **Total RTI consults.** | **372,783** | **172,468** | **-53.7** |

**Table S3.2:** **Change in azithromycin prescribing (Px) in all RTI consultations**

| | All RTI Consultations in RSC | | % Change |
|---|---|---|---|
| | **2019** | **2020** | *p<0.0001* |
| RTI no doxycycline Px | 328580 | 145232 | -55.8 |
| RTI with doxycycline Px | 44203 | 27236 | -38.4 |
| **Total RTI consults.** | **372,783** | **172,468** | **-53.7** |

**Table S3.3:** **Change in doxycycline prescribing (Px) in all RTI consultations**

We next have compared the change between years in consultations where azithromycin or doxycycline were prescribed (Table S3.4)

| | RTIs in RSC prescribed azithromycin | | % Change | RTIs in RSC prescribed doxycycline | | % Change |
|---|---|---|---|---|---|---|
| | **2019** | **2020** | | **2019** | **2020** | |
| **LRTI** | 1208 | 788 | -34.8 | 30318 | 15599 | -48.5 |
| **URTI** | 659 | 399 | -39.5 | 13357 | 10148 | -24.0 |
| **ILI** | 31 | 61 | 96.8 | 528 | 1489 | 182.0 |
| **Total** | **1898** | **1248** | **-34.2** | **44203** | **27236** | **-38.4** |

**Table S3.4:** **Change in prescribing pattern 2019 and 2020 in consultations where an antibiotic was prescribed**

These data allow us to report the change in the percentage of people with RTI prescribed our antibiotics of interest, azithromycin and doxycycline, and the change between years (Table S3.5).

| **Overall % RTI consultations** | **2019** | **2020** | **% change** |
|---|---|---|---|
| Prescribed azithromycin | 0.51 | 0.72 | 42.1 |
| Prescribed doxycycline | 11.86 | 15.79 | 33.1 |

**Table S3.5:** **Change in the proportion of RTI consultations in which our antibiotics of interest were prescribed**

**Negative Binomial Models for Doxycycline**

These models compare 2019 prescribing with 2020 (Table S3.6) and then look at whether those with COVID-19 are more likely to be prescribed doxycycline (Table S3.7).

| Doxy. Px Incidence Rates | IRR | Lower 95% CI | Upper 95% CI | p |
|---|---|---|---|---|
| **Yr 2020 (ref level 2019)** | 1.012 | 0.994 | 1.030 | 0.199 |
| **Age Band (ref. level 0-15)** | | | | |
| *16-64* | 12.34 | 11.86 | 12.85 | <0.0001 |
| *65+* | 45.46 | 43.53 | 47.48 | <0.0001 |
| **Gender (ref. level F)** | 0.79 | 0.78 | 0.81 | <0.0001 |
| **IMD Quintile (ref. level Most Deprived)** | | | | |
| *Q2* | 0.89 | 0.87 | 0.91 | <0.0001 |
| *Q3* | 0.84 | 0.81 | 0.86 | <0.0001 |
| *Q4* | 0.82 | 0.79 | 0.84 | <0.0001 |
| *Q5 (least deprived)* | 0.71 | 0.69 | 0.73 | <0.0001 |
| **NHS Region (Ref London)** | | | | |
| *Midlands and East* | 1.71 | 1.66 | 1.75 | <0.0001 |
| *North East and Yorkshire* | 1.55 | 1.50 | 1.60 | <0.0001 |
| *North West* | 1.28 | 1.24 | 1.32 | <0.0001 |
| *South East* | 1.11 | 1.08 | 1.14 | <0.0001 |
| *South West* | 1.60 | 1.55 | 1.65 | <0.0001 |
| **Resp. Disease** | | | | |
| *LRTI Count* | 1.0061 | 1.0056 | 1.0066 | <0.0001 |
| *URTICount* | 1.0003 | 1.0000 | 1.0007 | 0.0477 |
| *ILI Count* | 1.0170 | 1.0148 | 1.0193 | <0.0001 |

**Table S3.6:** **Model reporting the incident rate ratio (IRR) comparing prescribing of doxycycline in 2020 with 2019. Taking the variables in the model into account there was no difference in prescribing**

| Doxycycline presribing rate | IRR | Lower 95% CI | Upper 95% CI | p |
|---|---|---|---|---|
| **Covid19 Confirmed Count** | 1.0003 | 1.0002 | 1.0005 | 0.0001 |
| **Age Band (ref. level 0-15)** | | | | |
| *16-64* | 12.3 | 11.6 | 13.1 | <0.0001 |
| *65+* | 43.6 | 41.0 | 46.4 | <0.0001 |
| **Gender (ref. level F)** | 0.79 | 0.77 | 0.81 | <0.0001 |
| **IMD Quintile (ref. level Most Deprived)** | | | | |
| *Q2* | 0.88 | 0.85 | 0.92 | <0.0001 |
| *Q3* | 0.84 | 0.81 | 0.88 | <0.0001 |
| *Q4* | 0.82 | 0.79 | 0.85 | <0.0001 |
| *Q5 (least deprived)* | 0.71 | 0.69 | 0.74 | <0.0001 |
| **NHS Region (Ref London)** | | | | |
| *Midlands and East* | 1.69 | 1.63 | 1.76 | <0.0001 |
| *North East and Yorkshire* | 1.45 | 1.38 | 1.52 | <0.0001 |
| *North West* | 1.23 | 1.18 | 1.28 | <0.0001 |
| *South East* | 1.07 | 1.02 | 1.12 | 0.002 |
| *South West* | 1.52 | 1.46 | 1.59 | <0.0001 |
| **Resp. Disease** | | | | |
| *LRTI Count* | 1.0098 | 1.0088 | 1.0109 | <0.0001 |
| *URTI Count* | 1.0002 | 0.9995 | 1.0009 | 0.551 |
| *ILI Count* | 1.021 | 1.017 | 1.024 | <0.0001 |

**Table S3.7:** **Doxycycline prescribing in cases of COVID-19, for each unit rise in COVID-19 cases there has been a non-clinically significant rise in prescribing. Age 65 years and older, female gender, being more deprived, northern regions LRTI or ILI infections are all associated with a higher rate of prescribing**

**Comparison with Open Prescribing:**

We compared the differences in the rate of prescribing in OpenPrescribing between 2019 and 2020 with those in the RSC. The increase in azithromycin prescribing was almost identical. Both datasets showed a decrease in doxycycline prescribing however it was -2.31% in OpenPrescribing compared with -7.02% in the RSC (Table S4.1).

| | **2019** | **OpenPrescribing** | | **2020** | **OpenPrescribing** | | **2019** | **RCGP** | | **2020** | **RCGP** |
|---|---|---|---|---|---|---|---|---|---|---|---|
| | **Doxy** | **Azith** | | **Doxy** | **Axith** | | **Doxy** | **Azith** | | **Doxy** | **Axith** |
| 01/01/2019 | 330019 | 67232 | 01/01/2020 | 357980 | 71487 | 01/01/2019 | 20059 | 4189 | 01/01/2020 | 21389 | 4356 |
| 01/02/2019 | 262533 | 60916 | 01/02/2020 | 273769 | 64557 | 01/02/2019 | 15827 | 3699 | 01/02/2020 | 16375 | 3852 |
| 01/03/2019 | 249507 | 64264 | 01/03/2020 | 321979 | 77963 | 01/03/2019 | 14818 | 3826 | 01/03/2020 | 21131 | 5240 |
| 01/04/2019 | 245028 | 61168 | 01/04/2020 | 276404 | 73153 | 01/04/2019 | 15120 | 3852 | 01/04/2020 | 16295 | 4087 |
| 01/05/2019 | 231681 | 63809 | 01/05/2020 | 197299 | 64352 | 01/05/2019 | 14095 | 3762 | 01/05/2020 | 11810 | 3723 |
| 01/06/2019 | 206910 | 58523 | 01/06/2020 | 178034 | 64237 | 01/06/2019 | 12412 | 3342 | 01/06/2020 | 11051 | 3938 |
| 01/07/2019 | 206731 | 59961 | 01/07/2020 | 175987 | 65058 | 01/07/2019 | 13156 | 3853 | 01/07/2020 | 10930 | 3975 |
| 01/08/2019 | 201024 | 62694 | 01/08/2020 | 156884 | 58762 | 01/08/2019 | 11553 | 3522 | 01/08/2020 | 9503 | 3560 |
| 01/09/2019 | 211819 | 61199 | 01/09/2020 | 205472 | 65440 | 01/09/2019 | 13160 | 3791 | 01/09/2020 | 12755 | 4145 |
| 01/10/2019 | 275028 | 68038 | 01/10/2020 | 220466 | 68308 | 01/10/2019 | 16807 | 4124 | 01/10/2020 | 13463 | 4140 |
| **Total 2019** | **2420280** | **627804** | **Total 2020** | **2364274** | **673317** | 01/11/2019 | 17008 | 4020 | 01/11/2020 | 12676 | 4177 |
| *N.B. Open prescribing data only availabe for 10months in 2020* | | | | | | 01/12/2019 | 20079 | 4382 | 01/12/2020 | 13789 | 4405 |
| | | | | | | **Total 2019** | **184094** | **46362** | **Total 2020** | **171167** | **49598** |
| ***Difference between years*** | | | | -5600.6 | 4551.3 | ***Difference between years*** | | | | -12927 | 3236 |
| ***% Difference between years*** | | | | **-2.31** | **7.25** | ***% Difference between years*** | | | | -7.02 | 6.98 |

**Table S4.1 Comparison of prescribing of azithromycin and doxycycline in 2019 and 2020 in OpenPrescribing (national dataset, but data only to October 2020) and RSC. In both azithromycin prescribing increased by 7%. In both doxycycline prescribing reduced, but by 2.31 in OpenPrescribing and 7% in the RSC**
